# Associations of Systemic Inflammation and Senescent Cell Biomarkers with Clinical Outcomes in Older Adults with Schizophrenia

**DOI:** 10.1101/2024.03.06.24303857

**Authors:** M.K. Kirsten Chui, Kevin Schneider, Katherine Miclau, Sara C LaHue, David Furman, Heather Leutwyler, John C Newman

## Abstract

Individuals with schizophrenia suffer from higher morbidity and mortality throughout life partly due to acceleration of aging-related diseases and conditions. Systemic inflammation is a hallmark of aging and is also observed in schizophrenia. An improved understanding of how inflammation and accelerated aging contribute to long-term health outcomes in schizophrenia could provide more effective treatments to preserve long-term cognitive and physical function. In this pilot cross-sectional study, 24 older adults (≥55 years old) with schizophrenia were assessed on symptoms (Positive and Negative Syndrome Scale), neurocognition (Matrics Consensus Cognitive Battery), mobility (Timed Get Up and Go), and general health (SF-12). Serum levels of 112 different cytokines were measured, from which we derived estimated senescence-associated secretory phenotype (SASP) scores for each participant. Two-tailed Pearson’s bivariate correlations were computed to test the associations between schizophrenia clinical outcomes with individual cytokines, and SASP. Higher levels of eotaxin, IL-1α, IL-1β, and IFNα are associated with both worse PANSS negative and depressive symptoms scores. IL-1α and IL-1β negatively associated with general physical health whereas eotaxin negatively associated with mobility and global cognition. Overall, we found that specific inflammatory cytokines, but not composite measurements of SASP, are associated with clinical outcomes in older adults with schizophrenia.

## Introduction

Schizophrenia is a severe psychiatric disorder characterized by delusions, hallucinations, depressive symptoms and disordered thoughts and behaviors, which affects approximately 24 million people worldwide^1^. It is a debilitating disease that brings substantial mental, physical, and financial burden to the affected individuals and their caregivers. For older adults living with schizophrenia, deteriorating health is accompanied by the ever-increasing health-care related cost as the patient ages; the annual medical-related expenditures for patients with schizophrenia over the age of 65 is estimated to be nearly $40,000, markedly higher than that for older adults with depression, dementia, or other medical disorders^2^. A growing body of research suggests that schizophrenia is a disease of accelerated aging. It has long been associated with an increased risk of developing, and dying from, age-related diseases at a younger age than those without the disease^3^. People with schizophrenia have a 20% reduced life expectancy^4^ and an all-cause mortality rate 2.5 times greater compared to age-matched individuals without schizophrenia^5,6^. While some of this morbidity and mortality is accounted for by suicide and accidents, much is instead due to earlier onset of chronic diseases and conditions of aging^3,7,8^. Understanding the biological mechanisms driving this phenomenon is a key research priority to improve the lives and reduce the care burden of people aging with schizophrenia^9–11^.

Many studies found cellular and molecular evidence of accelerated aging in schizophrenia patients. Telomere attrition is a feature of aging, and significantly shorter leukocyte telomere length is observed in patients with schizophrenia and related disorders when compared to age-matched healthy individuals ^12,13^. The free radical theory attributes the accumulation of oxidative damages in cells and tissues to the functional decline in aging^14^.

Schizophrenia patients are reported to have significantly higher levels of oxidative stress and DNA damage^15–17^. Systemic chronic inflammation^18^ is another hallmark of aging found in schizophrenia. C-reactive protein (CRP), IL-6, IL-18, sIL-2R and TNFα are some proinflammatory cytokines reported to be elevated in schizophrenia patients^19–21^. A similar sterile and chronic inflammatory state with elevated CRP, IL-6, IL-18, TNFα (and other factors) is also observed in the aging population, which is termed “inflammaging”. Inflammaging is a risk factor for and associated with many age-related conditions^22,23^. Taken together, such biological changes are probable contributors to the observed increased risked for age-related diseases and mortality in patients with schizophrenia.

A key question for assessing the causality of inflammaging in schizophrenia is whether elevated inflammatory markers are associated with negative clinical outcomes among people with schizophrenia, and not only with having schizophrenia versus not having schizophrenia. One study reported positive correlation between CRP and positive symptoms with a moderate effect size (r = 0.491)^15^. Another study showed that IL-6 and TNFα positively correlated with depressive symptoms (but not positive or negative symptoms), however, the association was no longer significant after adjusting for potential confounds such as anti-inflammatory medications and BMI^21^. A meta-analysis of 25 similar studies demonstrated significant negative correlation between inflammatory cytokine levels and cognitive function^24^. Most existing studies shared a common limitation where only a small number of cytokines were investigated. Given the heterogenous nature of both schizophrenia and inflammaging, a more comprehensive analysis might reveal potentially causal molecules, provide more precise quantification of inflammaging, or provide clues to underlying mechanistic pro-inflammatory pathways.

Many of the cytokines implicated in schizophrenia are components of the Senescence-Associated Secretory Phenotypes (SASP). SASP is comprised of cytokines and signaling molecules secreted by senescent cells, cells with various forms of damage that enter a state of permanent cell cycle arrest and accumulate with age^25^. Senescence and SASP have been implicated in many age-related diseases and senescence burden correlates with higher medical comorbidity and worse cognitive functions in individuals with late-life depression^26^. While schizophrenia is not a disease of aging, strong indications of accelerated aging together with the similar inflammatory/senescence profile seen in patients suggest the possibility of common mechanisms underlying both conditions. In this pilot study, we explore if measurements of inflammaging, including individual cytokines, and composite measures of SASP, correlate with important clinical outcomes in a clinically well-characterized, cross-sectional cohort of older individuals with schizophrenia. While no statistically significant correlations were observed with SASP, we identified novel associations between individual cytokines (eotaxin, IL-1α, IL-1β, and IFNα) with specific clinical outcomes.

## Methods

### Design

Cytokine analyses were run on serum collected from a cross-sectional study of adults aged 55 years or older with schizophrenia or schizoaffective disorder. The cohort has been previously described in detail^27,28^, but key elements of the cohort and clinical data collection methodology are provided here for clarity. Institutional review board approval (#10-00443) was obtained from the Sponsoring University’s Institutional Review Board. Anonymity and confidentiality were upheld according to ethical guidelines.

### Participants and Settings

Participants were recruited from three sites: a transitional residential and day treatment center for older adults with severe mental illness; a locked residential facility for adults with serious mental illness; and an intensive outpatient case-management program. We recruited from 3 sites to obtain a sample with a range of psychiatric symptoms. IRB-approved recruitment information about the study was posted at these sites, and staff were also informed about the study. Participants were also referred from the community and other participants.

Remuneration consisted of $60 after completion of the first assessment, $30 after wearing a physical-activity monitor for 1 week, and a bonus payment of $20 all study procedures were completed. Inclusion criteria were: English-speaking adults older, aged 55 years or older, with a DSM-IV diagnosis of schizophrenia or schizoaffective disorder who passed a capacity-to-consent test based on comprehension of the consent form.

### Procedures

Data collection started in May 2010 and concluded in July 2012. Assessments took place at a private meeting room. Assessments included completion of a demographic questionnaire, confirmation of diagnosis using the Structured Clinical Interview for DSM Disorders (SCID), neurocognitive assessments, and assessment of schizophrenia symptoms. Additional clinical data obtained by research staff were blood pressure, waist circumference, weight, height, and patient report of smoking status and educational level. Typically, it took two to five meetings to complete the assessment.

### Measures

#### A trained member of the research staff administered study assessments

Symptomatology—Psychiatric symptoms were assessed with the extended Positive and Negative Syndrome Scale (PANSS)^29^. The extended PANSS contains six subscales measuring positive (e.g. delusions and hallucinations), negative (e.g. emotional and social withdrawal), excited (e.g. poor impulse control), depressed-anxious (e.g. depressed), disorganized (conceptual disorganization) and other symptoms (e.g. preoccupation, poor attention). The scale has demonstrated good-to-excellent reliability in assessing symptoms and their change during the course of treatment in clinical trials with participants diagnosed with schizophrenia^30^. The items on the PANSS are summed for the six subscales and the total score (the sum of all six subscales). The scale takes approximately 60 minutes to administer. PANSS scores were then converted to standardized z-score for statistical analyses.

Neurocognition: We used the Matrics Consensus Cognitive Battery (MCCB) to assess neurocognitive functioning on each of the following seven domains identified as discrete, fundamental dimensions of cognitive impairment in schizophrenia^31,32^: Speed of processing (SOP), Attention/ vigilance, working memory, verbal learning, visual learning, reasoning and problem solving, and social cognition. SOP tests include: Trail Making Test: Part A, Brief Assessment of Cognition in Schizophrenia (BACS) Symbol Coding, and Category Fluency (animal naming). Attention/Vigilance tests consisted of Continuous Performance Test-Identical Pairs (CPT-IP). Working memory tests used: Wechsler Memory Scale (WMS) -Spatial span and letter number span. The verbal learning test was the Hopkins verbal learning test revised. Visual Learning measures were the Brief Visuospatial Memory Test-revised. Reasoning and problem-solving measures consisted of Neuropsychology Assessment Battery (NAB) Mazes. Social Cognition included the Mayer-Salovey-Caruso Emotional Intelligence Test: Managing Emotions.

The MCCB scoring program yields seven domain scores and a composite score that are standardized to the same t-score measurement scale with a mean of 50 and an SD of 10 based upon the normative data collected from a sample of 300 community participants as part of the MATRICS psychometric and standardization study and published in the MCCB manual and the MCCB scoring program ^31–33^.

The SF-12 is a general measure of health status and was developed to be a shorter yet valid alternative to the SF-36^34^. The SF-12 includes one or two items from each of the following health domains: Physical functioning, role limitations due to physical health problems, bodily pain, general health, vitality (energy/fatigue), social functioning, role limitations due to emotional problems, and mental health (psychological distress and psychological well-being). It takes about 2 minutes to administer the SF-12. A score that ranges from 0 to 100 is generated, with higher scores indicative of better health status^35^. Then, these scores are standardized for the general population. For example, a score of 50 reflects an average level of physical health, and each 10-point change in score reflects 1 standard deviation.

Mobility—We assessed mobility objectively with the Timed Get Up and Go (TGUG) test. The TGUG is a valid and reliable test for quantifying functional mobility and is useful in following clinical change over time^36^. The test is easy to administer, quick and requires no special equipment. For the test, the person is observed and timed while they rise from an armchair, walks 3 meters, turns, walks back, and sits down again. A time of 12 seconds or less to complete the TGUG is used to identify normal mobility in community-dwelling adults and to differentiate fallers from non-fallers^37,38^. Standardized z-scores were reported.

### Cytokine Profiling

Serum samples were profiled for cytokines and chemokines using the Human Cytokine/Chemokine 96-Plex Discovery Assay Array (HD96), the Human Supplemental Biomarker 10-Plex Discovery Assay Array (HDHSB10), and the Human Cardiovascular Disease Panel 3 9-Plex Discovery Assay Array (HDCVD9), LASER Bead based multiplexing by Eve Technologies Corporation, Calgary, AB Canada. Each sample on each panel was run in singlet at 1:1, 1:100, and 1:40000 dilution respectively. The mean fluorescence intensity MFI values were used for all the statistical calculations. If the same analyte is analyzed on more than 1 panel, measurement from the one with better sensitivity is used.

### Correlation Analyses with Individual Cytokines

Cytokine MFI values were log-transformed and converted to z-scores. Two-tailed Pearson correlation coefficients at 95% confidence interval were computed for each clinical variables and each of the following 22 cytokines: CRP, MIG/CXCL9, IP-10/CXCL10, eotaxin, GM-CSF, GROα, MCP-1, MCP-4, MIP-1α, MIP-1β, MIP-3α, IFNα, IFNγ, IL-1α, IL-1β, IL-6, IL-8, TNF, TRAIL, ICAM-1, VCAM-1, and VEGF, using GraphPad Prism 10.1.1. We focused on these cytokines as they are either known to be elevated in schizophrenia, have been implicated in inflammaging, and/or identified as SASP factors. While not exhaustive, this exploratory analysis allowed us to survey potential associations between specific cytokines and clinical outcomes. Due to the exploratory nature of this small pilot study, the reported p-values are unadjusted. The complete correlation analysis result for all variables can be found in Supplementary Table 1.

### Mean z-scores for SASP Components

Log-transformed MFI values for each cytokine were converted to z-scores and the mean z-score was calculated by averaging all the respective SASP components listed below. Our data included the 11/22 comprising SASP by Diniz et al.^39^, 35/125 comprising SenMayo^40^, and 36/62 comprising the Translational Geroscience Network (TGN) SASP panel.

Diniz et al.: MCP-1, MCP-4, MIP-1α, MIP-3α, CCL4, GM-CSF, GROα, IL-1β, IL-6, IL-8, and ICAM-1 SenMayo: CCL1, CCL2/MCP-1, CCL3/MIP-1A, CCL4/MIP-1B, CCL5/RANTES, CCL7/MCP-3, CCL8/MCP-2, CCL13/MCP-4, CCL20/MIP-3a, CCL24/Eotaxin2, CCL26/Eotaxin3, CSF1/M-CSF, CSF2/GM-CSF, CXCL1/GROa, CXCL8/IL-8, CXCL10/IP-10, CXCL12/SDF-1, CXCL16, EGF, Fas, FGF2, HMGB1, ICAM-1, IL-1α, IL-1β, IL-2, IL-6, IL-7, IL-10, IL-13, IL-15, IL-18, Serpine1/PAI-1, TNF, VEGF/VEGFA TGN SASP: Eotaxin, TARC, MIP-3α, MIP-3β, MCP-1, MDC, RANTES, CRP, GM-CSF, Fractalkine, GROα, IL-1α, IL-1β, IL-1ra, IL-2, IL-5, IL-6, IL-7, IL-8, IL-10, IL-13, IL-15, IL-17, EGF, Fas, FGF2, ICAM-1, IFNα, IFNγ, MPO, PDGF-AA, PDGF-AB/BB, PAI-1, TNF, VCAM-1, and VEGF

## Results

A total of 24 participants (mean age 59.5, sd = 3.4) were included in the analyses. Sociodemographic and clinical measurements are shown in Table 1 and 2 respectively. The average total PANSS score was 77.1 with a standard deviation of 23.1, corresponding to “moderately ill” in Clinical Global Impressions (CGI)^41^. The mean MCCB global overall composite T-score was 18.9 (sd = 14.6, range = −14 to 42).

**Table 1.** Participants Sociodemographic Characteristics.

| Variable | Mean (SD) | Percent | N |
| --- | --- | --- | --- |
| Age, years | 59.5 (3.4) |  | 24 |
| Education, years | 13.1 (3.1) |  | 22 |
| Smoking years | 26.2 (19.0) |  | 24 |
| Smoker |  |  | 24 |
| Current |  | 58.3% |  |
| Past |  | 20.8% |  |
| Never |  | 20.8% |  |
| Male |  | 70.8% | 24 |
| Employed |  | 27.3% | 22 |
| Stable Residence |  | 66.7% | 21 |

**Table 2.** Mean (SD) Scores of Clinical Measurements (N = 24). ^a^N = 23.

| SF-12 (general health status) |  |
| --- | --- |
| Physical Health Summary | 42.1 (8.8) |
| Mental Health Summary | 44.5 (8.8) |
| TGUGT (mobility) | 11.6 (4.4) |
| PANSS (psychiatric symptoms) |  |
| PANSS total<sup>a</sup> | 77.1 (23.1) |
| PANSS negative | 14.1 (5.4) |
| PANSS depressed/anxious | 10.3 (3.7) |
| MCCB (global neurocognition) | 18.9 (14.6) |

First, we tested the association of individual cytokines with age and key clinical variables. We utilized measurements of general physical (SF-12) and cognitive (MCCB) health, as well as mobility (TGUGT, timed get up and go test) which capture the overall aging-related health and function of our participants. We also focused on PANSS negative and depressive symptoms as these are more strongly associated with long-term clinical outcomes and more treatment resistant than positive symptoms in older individuals with schizophrenia^42,43^. A heat map showing the associations between individual cytokines and clinical measurements is presented in Figure 1.

**Figure 1.**
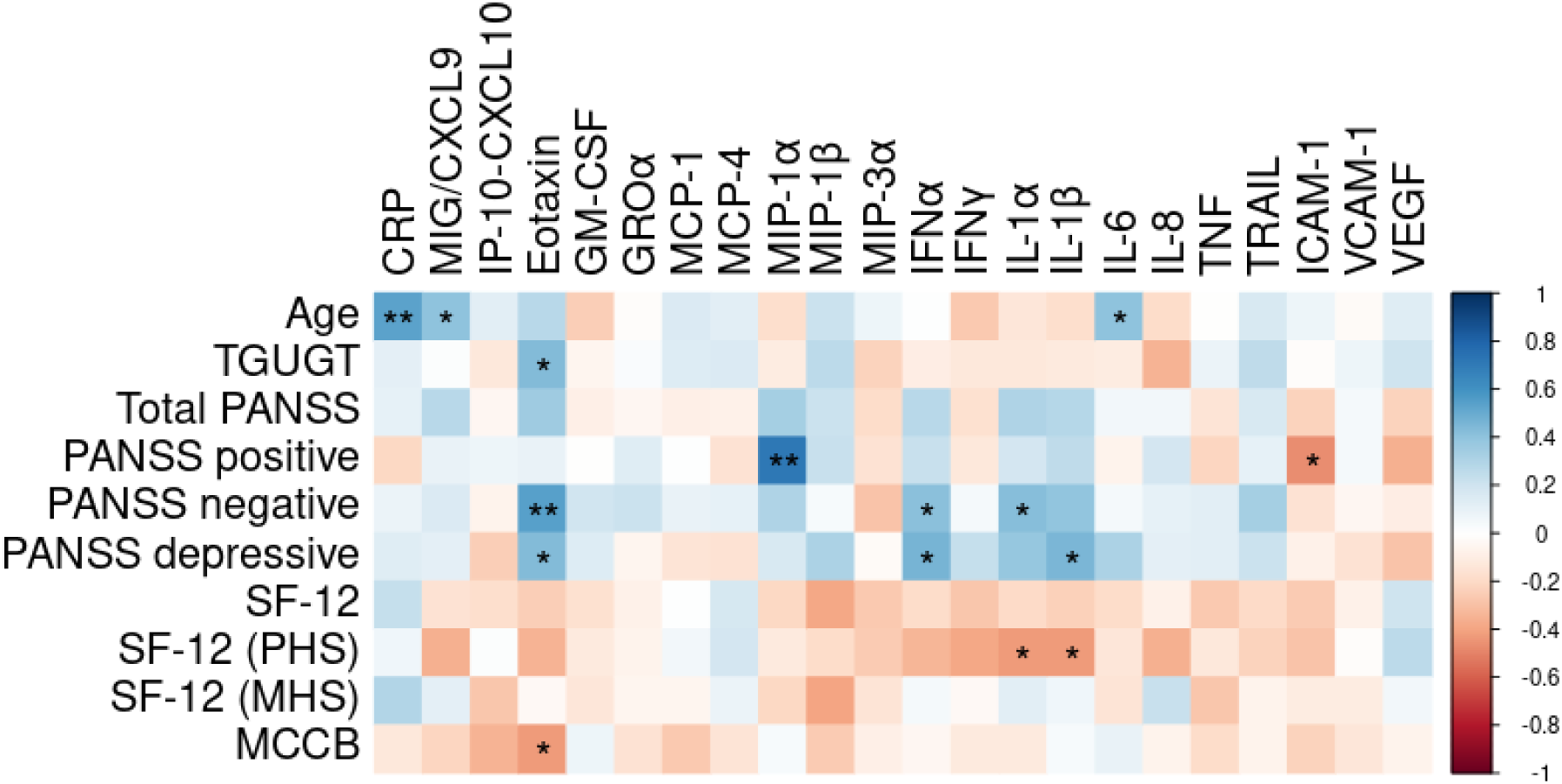
Heat Map Representation of Associations between Clinical Measurements and the 22 Cytokines Analyzed. Among the common inflammaging and SASP biomarkers investigated, many demonstrated positive association (in blue) with schizophrenia severity measured by PANSS. Many are also negatively associated (in red) with general health (SF-12) and global cognition (MCCB).

Major cytokines with significant clinical outcome associations are shown in Figure 2 and results from the correlation analyses for all the cytokines analyzed can be found in Supplementary Table 1. Due to the exploratory nature of this small pilot study, all reported p-values are unadjusted. Two-tailed Pearson correlation analyses demonstrated significant positive associations of CRP, MIG/CXCL9 and IL-6 with age (Fig. 2a), all of which have been implicated in inflammaging^22,23^. We observed that eotaxin, IL-1α, IL-1β, and IFNα levels are positively associated with schizophrenia severity as measured by PANSS. Higher serum levels of these cytokines are associated with both worse PANSS negative symptoms (Fig. 2b) and depressive symptoms scores (Fig. 2c), but not with positive symptoms or total (positive + negative symptoms) PANSS. IL-1α and IL-1β also negatively correlate with physical health measured by SF-12 (Fig. 2d). Higher levels of eotaxin are associated with higher TGUGT (reduced mobility) (Fig. 2e) and worse global cognitive function (MCCB) (Fig. 2f). Interestingly, while inflammaging can be observed in schizophrenia patients, the age-related markers (CRP, MIG/CXCL9, IL-6) are not associated with physical and cognitive health measures in these patients (Fig. 1 and Supp. Table 1). Our findings suggested that a distinct group of cytokines (eotaxin, IL-1α, IL-1β, and IFNα) may better represent clinical function.

**Figure 2.**
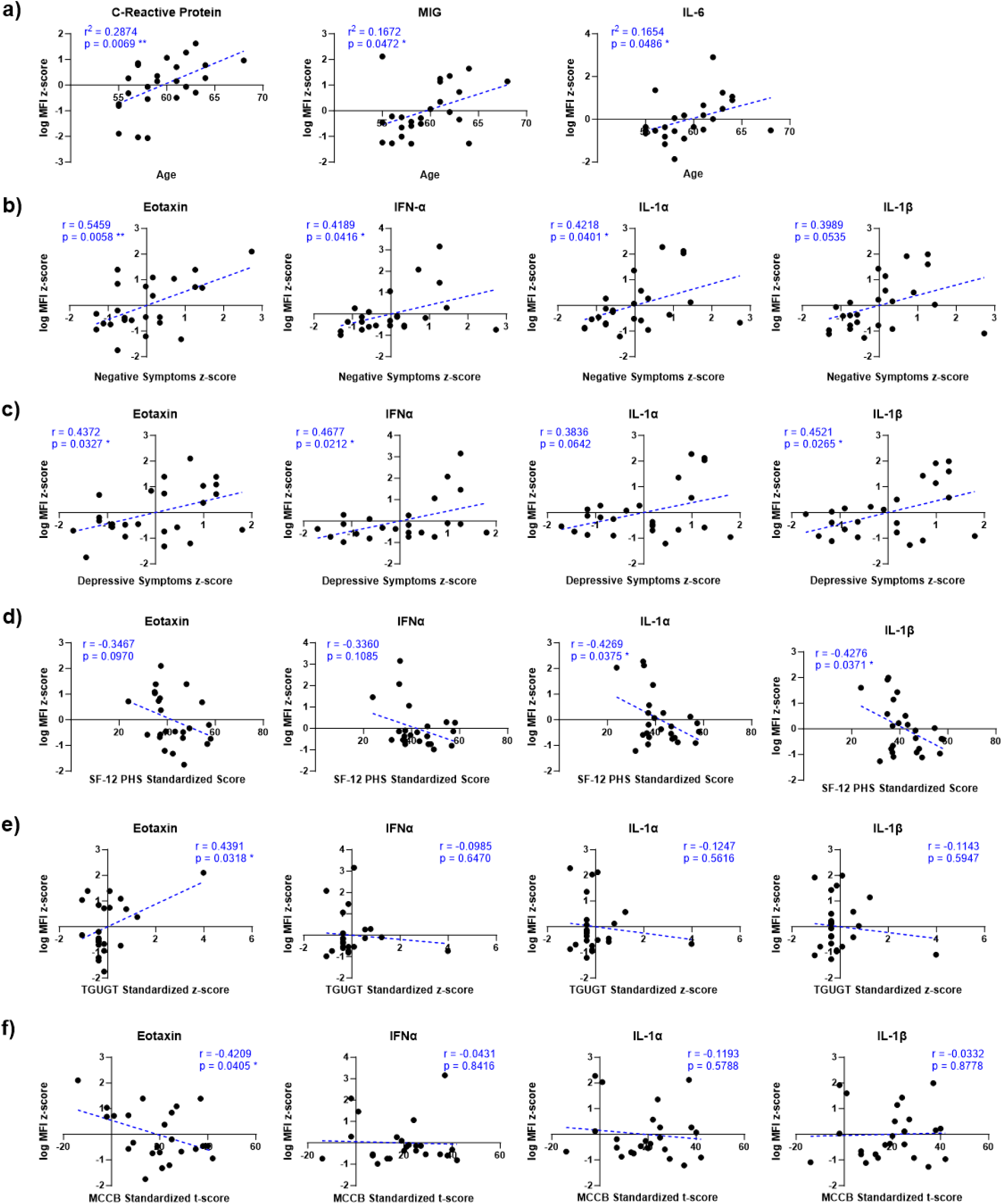
Relationship between Individual Cytokines, Age, and Clinical Measurements. (a) CRP, MIG, and IL-6 showed positive correlation with age. (b-c) Higher levels of eotaxin, IFNα, IL-1α, and IL-1β are associated with more PANSS negative and depressive symptoms. (d) IL-1α and IL-1β negatively associated with SF-12 physical health score. (e-f) Eotaxin also associated with reduced mobility and global cognition measured by TGUG test and MCCB respectively.

We then asked if cellular senescence, a specific mechanism underlying aging pathophysiology, may contribute to clinical outcomes in schizophrenia. While many SASP components have been identified, there is no standardized metric for quantifying senescence. Efforts have been made to generate SASP indices that estimate senescence in either specific clinical conditions such as major depression (by Diniz et al.^39^) or general aging (e.g. the SenMayo gene set^40^ and the Translational Geroscience Network SASP panel). We therefore explored how these SASP indices associate with SDoHs and symptoms severity in schizophrenia. Specifically, we asked if the mean level of the cytokines included in each of these indices which were also present in our data (see Methods) associated with clinical variables in our cohort. This is accomplished by calculating an unweighted mean z-score of the individual cytokine components of each index. We found consistent but non-significant associations between higher SASP and poorer physical, but not mental, health function (assessed by SF-12) (Fig. 3). No significant associations were observed with age, smoking, schizophrenia severity or global cognition (Table 3).

**Figure 3.**
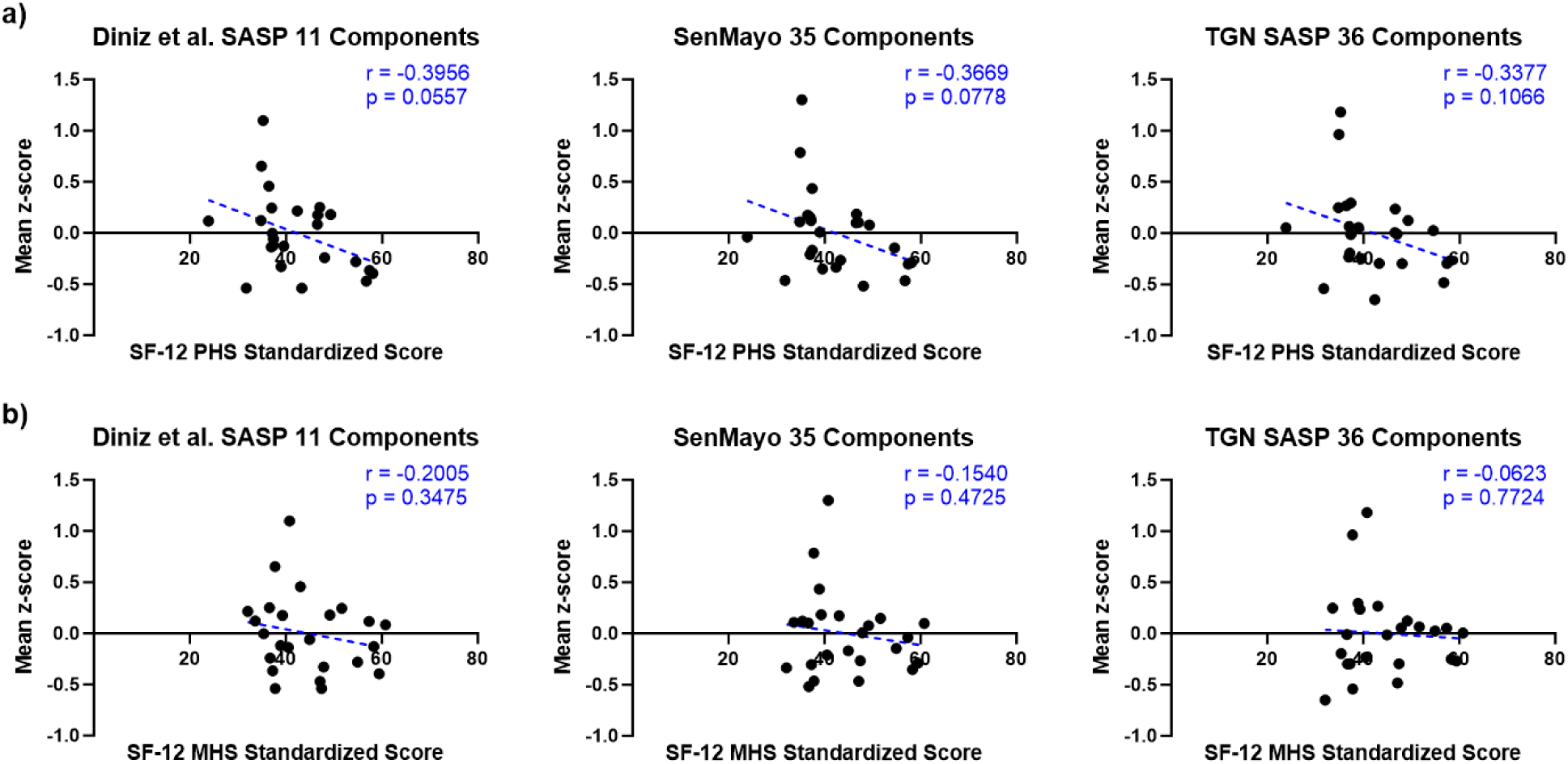
SASP Indices Components Correlation with SF-12 Global Health Functions. (a) Diniz et al. SASP, SenMayo, and TGN SASP components all demonstrated weak, non-significant, negative association with SF-12 PHS. (b) No apparent association observed between SASP and SF-12 MHS.

**Table 3.**
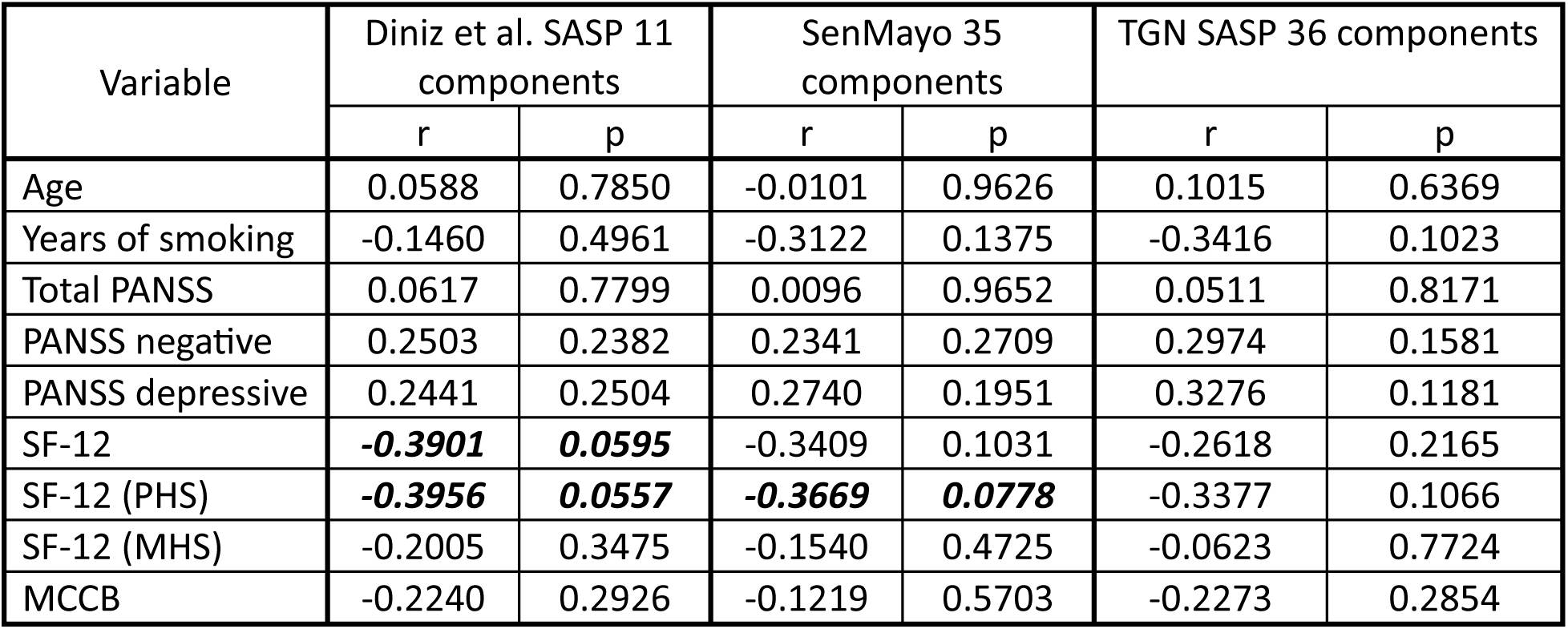
Summary of the Correlation Analyses with SASP components Mean Z-scores. Two-tailed Pearson correlation coefficients at 95% confidence interval were computed, Pearson r and unadjusted p-values reported in this summary table. Possible correlation trending toward significant (p<0.1) is italicized.

## Discussion

This study provides a comprehensive cytokine analysis of a small, well-characterized cohort of older adults with schizophrenia, including analysis of individual cytokines as well as composite SASP scores created to measure biological aging and underlying mechanisms of inflammaging. While our analyses are underpowered due to the small sample size relative to the large number of analytes, this pilot study is designed to survey potential cytokines and clocks of interest, which will guide future studies with larger, longitudinal, and potentially interventional cohorts.

Our findings of elevated levels of CRP, MIG/CXCL9, and IL-6 with age support previous evidence of inflammaging in schizophrenia. We also identified novel associations between specific cytokines (eotaxin, IL-1α, IL-1β, and IFNα) with general physical and cognitive health, mobility, and schizophrenia symptom severity. This was a pilot exploratory analysis, so individual cytokine results must be validated in larger studies. More detailed study is also required of the mechanisms by which these cytokines impact health and function in both clinical and preclinical models. If validated, such cytokines and inflammaging in general may provide important new potential treatment targets to improve health and function in adults aging with chronic schizophrenia, especially given the limitations of current antipsychotic medications on improving negative symptoms, cognitive function, and long-term aging phenotypes^44–47^.

Results from previous studies that interrogated inflammatory cytokines in schizophrenia are somewhat inconsistent. Two independent studies conducted by Herniman et al. and Lee et al. reported positive correlation between depressive symptoms and the levels of TNFα, and IL-6 ^21,48^. Another study by Dimitrov et al. revealed positive association between 9 cytokines (including IL-1β and IL-6) and PANSS positive as well as total scores^49^. Interestingly, they found no relation between any cytokine and PANSS negative or depressive symptoms^49^. The discrepancy between these reports and our current study may be attributed to some key differences in our cohorts of participant and methods of assessment. The participants in the former two studies used different tools for depression assessment; they also had a mean age of 48.1^21^ and 25.6^48^ respectively, considerably younger than our participants with a mean age of 59.5. Dimitrov et al. utilized the same PANSS scale and had participants at more similar age. However, all their participants were under antipsychotic treatment and 53.2% were on anti-depressants^49^, in contrast to at least 18.2% of our participants were not taking antipsychotics and over 63.6% were taking antidepressant. A larger proportion of our cohort experienced past major substance abuse (75.0% vs 53.2%) and some were taking other medications such as benzodiazepines or mood stabilizers at the time of the study (related data not reported in the study by Dimitrov et al.). Differences in medications may affect cognitive/functional outcomes as well as systemic inflammation. Taken together, ours and others’ findings pointed to a need for more in-depth investigation to delineate how chronological aging, and medications affect global inflammation, and health in schizophrenia patients.

CRP and IL-6 are both well-established markers for inflammation known to be upregulated with age^22,23^. CRP is part of the innate immune system and participate in the classical complement pathway^50^, whereas IL-6 mediates energy metabolism during acute inflammation^51^. MIG/CXCL9 mechanistically contributes to aging phenotypes by regulating inflammation and proliferation in a human cell model of the aging endothelium^52^. Positive association of CRP, IL-6, and MIG/CXCL9 levels with age in schizophrenic individuals suggest the same driving force for inflammaging may also be responsible for the schizophrenia-related aging acceleration. In accordance with our findings, prior reports showed evidence of increased eotaxin in schizophrenia^53,54^, which is associated with worse negative symptoms^54^. Eotaxin (or CCL-11) has been reported to increase with age, is associated with risk of Alzheimer’s and other neurodegenerative diseases, and has been informally labeled as “Accelerated Brain-Aging Chemokine”^55^. Both IL-1α and IL-1β are proinflammatory components of the innate immune response^56^ that have been previously implicated in schizophrenia. However, there are contradicting reports regarding how the level of IL-1α changes in schizophrenia^57–59^. IL-1β was found to be elevated in patients during first-episode psychosis^60^ and in chronic schizophrenia^61^. Although our results agree with existing publications that IL-1α and IL-1β are important factors in the disease, more detailed studies are required to clarify the role they play in pathology. IFNα is a type I interferons classically known for their role in viral infections^62^. Information regarding how IFNα affect schizophrenia patients is limited, one longitudinal study showed that for non-responders to pharmacotherapy, higher initial IFNα level negatively correlate with PANSS improvement after clozapine treatment^63^. Taken together with our observations of positive correlations between IFNα level and PANSS negative and depressive symptoms, current indications suggest that IFNα negatively impact symptoms burden. IFNα may be targeted in future study as a possible intervention to manage schizophrenia symptoms.

Although eotaxin, IL-1α, IL-1β, and IFNα are also recognized as common SASP components, our results suggested that senescence and SASP may not explain inflammaging and schizophrenia symptoms in this clinical cohort. We found potential associations between SASP and global physical functions (SF-12 PHS) that did not reach statistical significance. However, we found no association between SASP score and symptom severity or cognitive function. Senescence is a new translational biology field, and to our knowledge there has been no study directly measuring senescent cell burden in older adults with schizophrenia. There is also not yet a standardized and validated definition of SASP for use in human studies. While senescent cell burden increases with age, and contributes to systemic chronic inflammation, it is possible that the biological and social stressors in schizophrenia generate chronic inflammation via different mechanisms or that senescence may have a role in physical but not mental or cognitive health in schizophrenia.

While there are a variety of reasons why people with schizophrenia experience accelerated aging, one important possibility is the cumulative effect of Social Determinants of Health (SDoH) adversity. SDoH are defined as the conditions in which people are born, grow, work, live, and age, and the wider set of social, political and economic forces shaping these conditions^64^. The presence and severity of psychosis has been associated with increased disadvantage for a number of SDoH, including exposure to early life adversity, social disconnection and fragmentation, housing instability, food insecurity, and incarceration^65^. SDoH adversity has been demonstrated to accelerate the aging process by promoting telomere attrition, oxidative stress, and systemic inflammation, which in turn leads to a range of age-related diseases^66,67^. Further study with a larger, longitudinal, interventional cohort is necessary to better understand how SDoH interrelate with molecular changes in aging and schizophrenia, which may elucidate mechanistic links. It will also be important to explore if interventions that reduce SDoH adversity, such as supportive housing, may reduce inflammaging in older individuals with schizophrenia.

## Limitations

There are limitations to our study, primarily due to the small size of the cohort, its cross-sectional nature, and the exploratory nature of our high-dimensional analyses. Due to the small sample size, we were not able to adjust for potentially important covariates such as age, sex, medications, and SDoH in the cytokine analyses. Since most of our participants were men, we are also limited in applying our results to women with schizophrenia. Our approach to estimate SASP had limitations beyond the lack of a validated SASP definition. For example, the SenMayo SASP list was originally generated and validated based on gene expression in tissue, rather than blood protein levels. Moreover, not all SASP factors within the three indices were present in our data. Finally, relative weighting and expected directionality for SASP factors in blood has not been established. Thus, our approach of calculating the mean z-score of available factors may not accurately represent SASP or senescent cell activity.

## Conclusion

In this pilot study we demonstrate associations between specific, distinct groups of cytokines with age and with clinical outcome measures, with a particularly notable role for eotaxin/CCL-11 in the latter. Cellular senescence, assessed via multicomponent SASP indicies, may not explain inflammaging and accelerated aging-related clinical outcomes seen in schizophrenia. Given the overwhelming evidence of accelerated aging in this condition, a better understanding of the role systemic inflammation and aging play in the pathophysiology of schizophrenia is paramount. Our findings build upon prior studies to emphasize the importance of systemic inflammation in schizophrenia. Further investigation in larger cohorts are necessary to define the molecular signatures that correspond to specific clinical outcomes, which may offer mechanistic insight into pathophysiology and more tailored treatment options.

## Data Availability

All data produced in the present study are available upon reasonable request to the corresponding authors.

## Funding

This work was supported by the UCSF Academic Senate [Individual Investigator Grant, H.L.]; National Center for Advancing Translational Sciences, National Institutes of Health through UCSF-CTSI [grant number KL2TR000143, H.L.]; the National Institute of Nursing Research [grant number P30-NR011934-0, H.L.], Buck Institute intramural funds (J.C.N.), and the Navigage Foundation (M.K.K.C.).

## Disclosures

D.F. is a co-founder of Edifice Health, a company that provides inflammation-based biomarker services. The other authors report they have no competing interests to declare.

## Notes

### Author Declarations

Institutional Review Board of the University of California, San Francisco gave ethical approval for this work.

